# The Cardiac Toxicity of Chloroquine or Hydroxychloroquine in COVID-19 Patients: A Systematic Review and Meta-regression Analysis

**DOI:** 10.1101/2020.06.16.20132878

**Authors:** Imad M. Tleyjeh, Zakariya Kashour, Oweida AlDosary, Muhammad Riaz, Haytham Tlayjeh, Musa A. Garbati, Rana Tleyjeh, Mouaz H. Al-Mallah, M. Rizwan Sohail, Dana Gerberi, Aref A. Bin Abdulhak, John R. Giudicessi, Michael J. Ackerman, Tarek Kashour

**Affiliations:** Infectious Diseases Section, Department of Medical Specialties King Fahad Medical City, Riyadh, Saudi Arabia; Division of Infectious Diseases, Mayo Clinic College of Medicine and Science, Rochester, MN, USA; Department of Epidemiology, Mayo Clinic College of Medicine and Science, Rochester, MN, USA; College of Medicine, Al Faisal University, Riyadh, Saudi Arabia; Department of Medicine, University of Alberta, Edmonton, Alberta, Ca; Department of Statistics, Quaid Azam University Islamabad, Pakistan; Department of Intensive Care, King Abdulaziz Medical City, King Saud bin Abdulaziz for Health Sciences and King Abdullah International Medical Research Center, Riyadh, Saudi Arabia; Infectious Diseases Unit, Department of Medicine, University of Maiduguri, Maiduguri, Nigeria; Houston Methodist DeBakey Heart and Vascular Center, Houston, TX, USA; Department of Cardiovascular Diseases, Mayo Clinic College of Medicine and Science, Rochester, MN, USA; Mayo Clinic Libraries, Mayo Clinic, Rochester, MN, USA; Swedish Heart and Vascular Institute, Swedish Medical Center, Seattle, WA, USA; Division of Heart Rhythm Services, Department of Cardiovascular Medicine; Department of Pediatric and Adolescent Medicine; Department of Molecular Pharmacology and Experimental Therapeutics, Mayo Clinic College of Medicine and Science, Rochester, MN, USA; Department of cardiac sciences, King Fahad Cardiac Center, King Saud University Medical City, Riyadh Saudi Arabia

## Abstract

**Importance:** The antimalarial agents chloroquine (CQ) and hydroxychloroquine (HCQ) have been proposed as a potential treatment for COVID-19 due their effect on several cellular processes that impact viral replication. Although more than 100 ongoing trials are testing their efficacy, CQ and HCQ are being used widely in clinical practice, exposing COVID-19 patients to potentially significant cardiac adverse effects.

**Objective:** To systematically review the literature and estimate the risk of cardiac toxicity in patients receiving CQ or HCQ for COVID-19.

**Data Sources:** A systematic search was conducted on May 27, 2020 of Ovid EBM Reviews, Ovid Embase (1974+), Ovid Medline (1946+ including epub ahead of print, in-process & other non-indexed citations), Scopus (1970+) and Web of Science (1975+) and preprint servers (Medrvix and ResearchSquare) and manual search of references lists.

**Study Selection:** Studies that included COVID-19 patients treated with CQ or HCQ, with or without azithromycin, were included as follows: (1) COVID-19 patient population, (2) the study included more than 10 patients receiving either one of the medications, (3) reported electrocardiographic changes and/or cardiac arrhythmias.

**Data Extraction and Synthesis:** Study characteristics and endpoints incidence were extracted. Due to the very low incidence of torsades de pointes (TdP) and other endpoints (rare events), the arcsine transformation was used to obtain a pooled estimate of the different incidences using a random-effects meta-analysis. Meta-regression analyses were used to assess whether the incidence of different endpoints significantly varied by multiple study-level variables specified a priori.

**Main Outcomes and Measures:** Pooled Incidence of: (1) change in QTc value from baseline ≥ 60 ms, (2) QTc ≥ 500 ms, (3) the composite of endpoint 1 and 2, (4) TdP arrhythmia or ventricular tachycardia (VT) or cardiac arrest, (5) discontinuation of treatment due to drug-induced QT prolongation or arrhythmias.

**Results:** A total of 19 studies with a total of 5652 patients were included. All included studies were of high methodological quality in terms of exposure ascertainment or outcome assessment. Among 2719 patients treated with CQ or HCQ, only two episodes of TdP were reported; the pooled incidence of TdP arrhythmia or VT or cardiac arrest was 3 per 1000, 95% CI (0-21), I^2^=96%, 18 studies with 3725 patients. Among 13 studies of 4334 patients, the pooled incidence of discontinuation of CQ or HCQ due to prolonged QTc or arrhythmias was 5%, 95% CI (1-11), I^2^=98%. The pooled incidence of change in QTc from baseline of ≥ 60 ms was 7%, 95% CI (3-14), I^2^=94% (12 studies of 2008 patients). The pooled incidence of QTc ≥ 500 ms was 6%, 95% CI (2-12), I^2^=95% (16 studies of 2317 patients). Among 11 studies of 3127 patients, the pooled incidence of change in QTc from baseline of ≥ 60 ms or QTc ≥ 500 ms was 9%, 95% CI (3-17), I^2^=97%. Mean/median age, coronary artery disease, hypertension, diabetes, concomitant QT prolonging medications, ICU care, and severity of illness in the study populations explained between-studies heterogeneity.

**Conclusions and Relevance:** Treatment of COVID-19 patients with CQ or HCQ is associated with a significant risk of drug-induced QT prolongation, which is a harbinger for drug-induced TdP/VT or cardiac arrest. CQ/HCQ use resulted in a relatively higher incidence of TdP as compared to drugs withdrawn from the market for this particular adverse effect. Therefore, these agents should be used only in the context of randomized clinical trials, in patients at low risk for drug-induced QT prolongation, with adequate safety monitoring.

**Key Points:** *Question:* What are the risks of different cardiac toxicities in patients receiving chloroquine (CQ) or hydroxychloroquine (HCQ) for COVID-19.

*Findings:* In this systematic review, treatment of COVID-19 patients with CQ or HCQ is associated with a clinically significant risk of drug-induced QT prolongation, and torsades de pointes (TdP) arrhythmia/ventricular tachycardia/cardiac arrest in a relatively higher incidence compared to drugs withdrawn from the market for such adverse effects.

*Meaning:* These agents should be used only in the context of clinical trials, in patients at low risk for drug-induced QT prolongation, with adequate safety monitoring.

## Introduction

The severe acute respiratory syndrome coronavirus 2 (SARS-CoV-2) that causes the coronavirus disease 2019 (COVID-19) has spread across the globe creating a significant impact on the public health by infecting millions of individuals and claiming hundreds of thousands of lives. The enormous death toll and economic losses produced by the pandemic along with the uncertainties about its treatment and prevention has generated significant public stress increasing the demand for developing effective therapies to control the spread of the disease. In the absence of proven and approved therapies and the significant time lag before an effective vaccine is available, repurposing of existing drugs for the treatment of COVID-19 disease may offer the fastest and most promising therapeutic approach.

Among the drugs that received significant early attention were the antimalarial agents chloroquine (CQ) and hydroxychloroquine (HCQ). CQ and HCQ are weak bases and diffuse easily across cell membranes to acidic cellular vesicles like lysosomes and late endosomes, raising their pH and thereby affecting several cellular processes like endocytosis, exosome release and phagolysosomal fusion^1^. These changes could affect several stages of viral life cycles from cell entry, viral replication, and viral particle assembly to viral particle release from the host cells^1^. CQ and HCQ also interfere with glycosylation process and this could alter angiotensin converting enzyme 2 (ACE2) processing and function and thereby could hinder the binding of SARS-CoV-2 virus to its receptor ACE2^1^.

Additionally, these vesicle pH changes modulate the immune system by interfering with antigen processing and presentation and consequently immune cell activation and production of proinflammatory cytokines like IL-6 and TNF-α1. This effect is favorable in the management of autoimmune diseases like systemic lupus erythematosus (SLE) and may have favorable impact in COVID-19 patients with cytokine storm. Among other beneficial effects of CQ and HCQ is their effect on vascular endothelial cells. It has been reported that HCQ prevents endothelial dysfunction through activation of endothelial nitric oxide (eNOS), reduction of inflammation, and reversal of prothrombotic state^2^. This observation is very pertinent to patients with COVID-19 since they manifest significant pulmonary vascular endothelialitis and microvascular thrombosis^3^.

The successful demonstration of in vitro antiviral properties of CQ and HCQ against SARS-CoV-2 virus led to initiation of several clinical studies testing the therapeutic potential of CQ and HCQ in COVID-19 ^4,5^. The early experience came from China where a report of 100 patients treated with CQ claimed to result in less severe pneumonia associated with improved pulmonary imaging and shorter disease course ^6^. Subsequent to this report, the National Health Commission of the People’s Republic of China released expert consensus recommending high dose of CQ of 500 mg twice daily for 10 days in patients with COVID-19 disease^7^. Another early non-randomized study of 20 COVID-19 patients treated with HCQ alone or in combination with azithromycin in France showed reduced nasopharyngeal viral carrier sate at 6 days after the initiation of treatment^8^. Despite its serious methodological limitations, this report received exceptional attention by media and politicians and triggered a widespread off label use of CQ and HCQ for COVID-19 with subsequent reports of CQ-related deaths^9^.

While CQ and HCQ are considered safe generally, QT prolongation and torsade de pointes (TdP) ventricular tachycardia as well as other arrhythmias, myocarditis, and cardiomyopathy have been reported^10-13^. The risk of drug-induced QT prolongation, drug-induced TdP, and other arrhythmias and structural complications could be exacerbated by coexisting morbidities like old age and cardiovascular conditions such as ischemic heart disease (IHD), heart failure, electrolyte imbalance, hypoxemia, and renal impairment. Several epidemiological studies observed higher prevalence of cardiovascular diseases like hypertension, IHD, and diabetes in patients with severe COVID-19^14-17^. Other investigations have revealed increased risk of arrhythmias, acute cardiac injury, myocarditis, heart failure, and thromboembolic complications associated with COVID-19^18-21^. This makes COVID-19 patients more vulnerable and susceptible to drug induced cardiac toxicities. Indeed, recent reports confirmed the increased risk of QT prolongation among COVID-19 patients^22,23^.

The indiscriminate use of the antimalarial medications for the treatment of COVID-19 in the absence of robust clinical evidence for their efficacy, coupled with associated potential harm, calls for rigorously conducted systematic reviews/meta-analyses of the available clinical data to present a clearer picture about their safety and efficacy and provide data-informed view regarding their utility in the treatment of COVID-19. In this study, we set to systematically review the literature regarding the cardiac toxicity of CQ or HCQ in patients with COVID-19.

## Methods

### Inclusion and Exclusion Criteria

We followed Preferred Reporting Items for Systematic Reviews and Meta-analyses (PRISMA)^24^ and Meta-Analysis of Observational Studies in Epidemiology (MOOSE)^25^ guidelines for reporting systematic review and meta-analysis of observational studies guidelines. Studies that reported electrocardiographic changes and/or cardiac arrhythmias in COVID-19 patients treated with CQ or HCQ with and without azithromycin were included using pre-specified inclusion criteria as follows: (1) COVID-19 patient population, (2) the study included more than 10 patients receiving either one of the agents, (3) electrocardiographic changes and/or cardiac arrhythmias were reported. In order to avoid introducing non-independence by including patients in the analysis more than once, we included results from the same study with the larger sample size if there were more than one study reporting data on overlapping patient population^22^.

### Literature Search

The literature was searched by a medical librarian for the concepts of CQ or HCQ combined with COVID-19. The search strategies were created using a combination of keywords and standardized index terms. Searches were run on May, 27, 2020 in Ovid EBM Reviews, Ovid Embase (1974+), Ovid Medline (1946+ including epub ahead of print, in-process & other non-indexed citations), Scopus(1970+) and Web of Science (1975+). Results were limited to 2019+. All results were exported to Endnote where obvious duplicates were removed. Search strategies are provided in the supplement 1. We also searched for unpublished manuscripts using the medRxiv services operated by Cold Spring Harbor Laboratory and Research Square preprints. In addition, we searched Google Scholar and the references of eligible studies and review articles. Two authors independently identified eligible studies (ZK, OA) and 4 authors (ZK, MG, OA, HT) extracted the data into a pre-specified data collection form. We collected data about study’s population and characteristics, and different cardiac toxicity endpoints. A senior author verified all data included in the analyses thrice (TK).

The eligible studies were assessed according to the Newcastle–Ottawa quality assessment scale (NOS) according to only 4 parameters relevant to our single arm meta-analysis by (HT, OA, ZK): exposure assessment, outcome assessment, length of follow up and loss to follow up rate ^26^.

### Study Endpoints

Endpoints included the incidence of: (1) change in QTc interval from baseline of ≥ 60 ms, (2) QTc ≥ 500 ms, (3) the composite of endpoint 1 and 2, (4) TdP arrhythmia, or ventricular tachycardia (VT) or cardiac arrest and (5) discontinuation of treatment due to prolonged QT or arrhythmias.

### Statistical Analysis

The number and percentage of subjects experiencing different endpoints were extracted from each study. Due to the very low incidence of TdP and other endpoints (rare events), the arcsine transformation was used to obtain a pooled estimate of the different incidences. For meta-analyses of very rare events, this transformation is more appropriate than the commonly used logit transformation, as it can accommodate studies with no observed events, without requiring a continuity correction^27-29^. Both fixed-effects and random-effects DerSimonian & Laird models with inverse variance method were considered^30^; the random-effects models were estimated using restricted maximum likelihood. Statistical tests for heterogeneity were performed using the Cochrane’s *Q* and I-squared statistics, with the assumption of homogeneity being considered invalid for *P* values < 0.05 ^31^. When homogeneity could not be assumed, we reported summary estimates and forest plots from the random-effects models.

Meta-regression analyses were used to assess whether the incidence of different endpoints significantly varied by multiple variables specified a priori. These variables were chosen based on risk factors that are known to potentially increase the risk of cardiac arrhythmias (age, sex, CAD, CHF, DM, disease severity, CKD, ICU care, and mortality as another surrogate of disease severity).

We then performed sensitivity analyses by repeating the above analyses after excluding two studies that used high dose CQ or HCQ. Two-tailed p values less than 0.05 were considered to be statistically significant. All statistical analyses were performed using meta and metafor packages in R statistical software, version 3.6.3.^32^

## Results

### Included Studies

A total of 19 studies with a total of 5652 patients, including single center and multicenter studies, from different countries were included in our systematic review^22,23,33-49^. Figure 1 shows the result of our search strategy (PRISMA flow diagram). Table 1 illustrates the general characteristics of the included studies. All included studies were of high methodological quality in terms of exposure ascertainment or outcome assessment (Table 2). Several important cardiovascular adverse events have been observed in our meta-analysis. However, several events have not been reported consistently by all of the included studies and therefore, we were unable to perform a meta-analysis for all the events.. The reported events that we could not meta-analyze were cardiac complications other than TdP, ventricular tachycardia and cardiac arrest.

**Table 1:** Characteristics of Included Studies

| Study | Country | Study population | Drugs used | Cardiac toxicity | Monitoring method |
| --- | --- | --- | --- | --- | --- |
| Tang et al | China | Hospitalized adult patients with mild to moderate or severe COVID-19 infection based on the 5th version of Chinese guidelines | HDQ 1200 mg daily for 3 days then 800 mg daily for 2 weeks or 3 weeks | QTc prolongation<br>Cardiac arrhythmia during therapy course | NR |
| Borba et al | Brazil | Hospitalized adult patients with suspected COVID and RR>24, HR>125, SpO2 <90% on RA or Shock state | High (total dose 12g) or low dose (total dose 2.7g) HDQ for 10 days<br>Azithromycin 500 mg for 5 days | QTc >500 ms or ventricular arrhythmia for 28 days | ECG on day 13,28<br>ECG done at clinician discretion |
| Perinel et al | France | Hospitalized critically ill patients | HDQ 200 mg 3 times daily for 5 days | NR | NR |
| Ramirredy et al | USA | Hospitalized patients with COVID-19 who were treated with azithromycin and/or HDQ | Azithromycin and /or HDQ | QT prolongation<br>Cardiac arrhythmia during therapy course | Daily ECG |
| Mahevas et al | France | Hospitalized patients with COVID-19 who received oxygen therapy | HDQ 600 mg daily. Duration NR | QTc prolongation<br>arrhythmia | Daily ECG until 5 days after drug discontinuation |
| Capriani et al | France | Hospitalized patients with COVID-19 who were treated with azithromycin and HDQ | HDQ 200 mg twice daily for at least 3 days<br>Azithromycin 500 mg daily | QTc prolongation<br>arrhythmia | Basal ECG and on day 3<br>Or 24 hour cardiac monitor for duration of therapy |
| Chorin et al | USA/Italy | Hospitalized patients with COVID-19 who were treated with azithromycin and HDQ | HDQ 400 mg twice daily day 1 then 200 mg twice daily for 4 days<br>Azithromycin 500 mg daily | QTc prolongation | Baseline ECG and at least one ECG after drug administration |
| Van Den Broek et al | Netherland | Hospitalized patients with COVID-19 who received respiratory | Chloroquine 600 mg daily for 5 days | QTc prolongation | Baseline ECG and at least one ECG after drug |

|  |  | support |  |  | administration |
| --- | --- | --- | --- | --- | --- |
| Saleh et al | USA | Hospitalized patients with COVID-19 who were treated with HDQ +/- azithromycin | HDQ +/- azithromycin<br>Dose/duration<br>NR | QTc<br>prolongation<br>arrythmia | Baseline ECG then twice daily ECG or mobile cardiac monitoring |
| Bessiere et al | France | Hospitalized patients with COVID-19 who were admitted to intensive care unit | HDQ 200 mg twice daily for 10 days +/- azithromycin 250 mg for 3 days | QTc<br>prolongation | Daily ECG<br>Continuous cardiac monitor |
| Chang et al | USA | Hospitalized patients with COVID-19 who were treated with HDQ +/- azithromycin | HDQ 400 mg twice daily day 1 then 200 mg twice daily for 4 days<br>+/- Azithromycin 500 mg daily | QTc<br>prolongation | Mobile cardiac monitor |
| Mercuro et al | USA | Hospitalized patients with COVID-19 who were treated with HDQ +/- azithromycin | HDQ 400 mg twice daily day 1 then 200 mg twice daily for 4 days | QTc<br>prolongation | ECG in electronic health records |
| Ip et al | USA | Hospitalized patients with COVID-19 who were treated with HDQ +/- azithromycin | HDQ, azithromycin or combination<br>Dose and duration NR | QTc<br>prolongation<br>arrythmia | ECG in electronic health records |
| Jain et al | USA | Hospitalized patients with COVID-19 | HDQ, dose and duration NR | QTc<br>prolongation | ECG or telemetry monitoring |
| Million et al | France | Inpatients and outpatients with COVID-19 | HDQ 600 mg daily for 10 days<br>Azithromycin 500 mg day 1 then 250 mg for 4 days | QTc<br>prolongation | Baseline ECG then ECG on day 2 of treatment |
| Molina et al | France | Hospitalized patients with COVID-19 | HDQ 600 mg daily for 10 days<br>Azithromycin 500 mg day 1 then 250 mg for 4 days | QTc<br>prolongation | NR |
| Pareira et al | USA | Hospitalized solid organ transplant patients with COVID-19 | HDQ 600 mg twice daily day 1 then 200 mg twice daily for 4 days<br>Azithromycin<br>500 mg day 1<br>then 250 mg for 4 days | QTc<br>prolongation | Baseline ECG<br>ECG on day 2 or 3 of therapy |
| Rosenberg et al | USA | Hospitalized patients with COVID-19 | HDQ and Azithromycin alone or as combination<br>Dose and duration NR | Abnormal ECG (arrhythmia or QT prolongation) | Random ECG screening |
| Ruiz et al | Spain | Hospitalized solid organ transplant patients with COVID-19 | HDQ 400 mg twice daily day 1 then 200 mg twice daily for 4 days | NR | NR |

Abbreviations: HDQ, hydroxychloroquine: ECG, electrocardiogram: NR, not reported: MS, millisecond

**Table 2:**
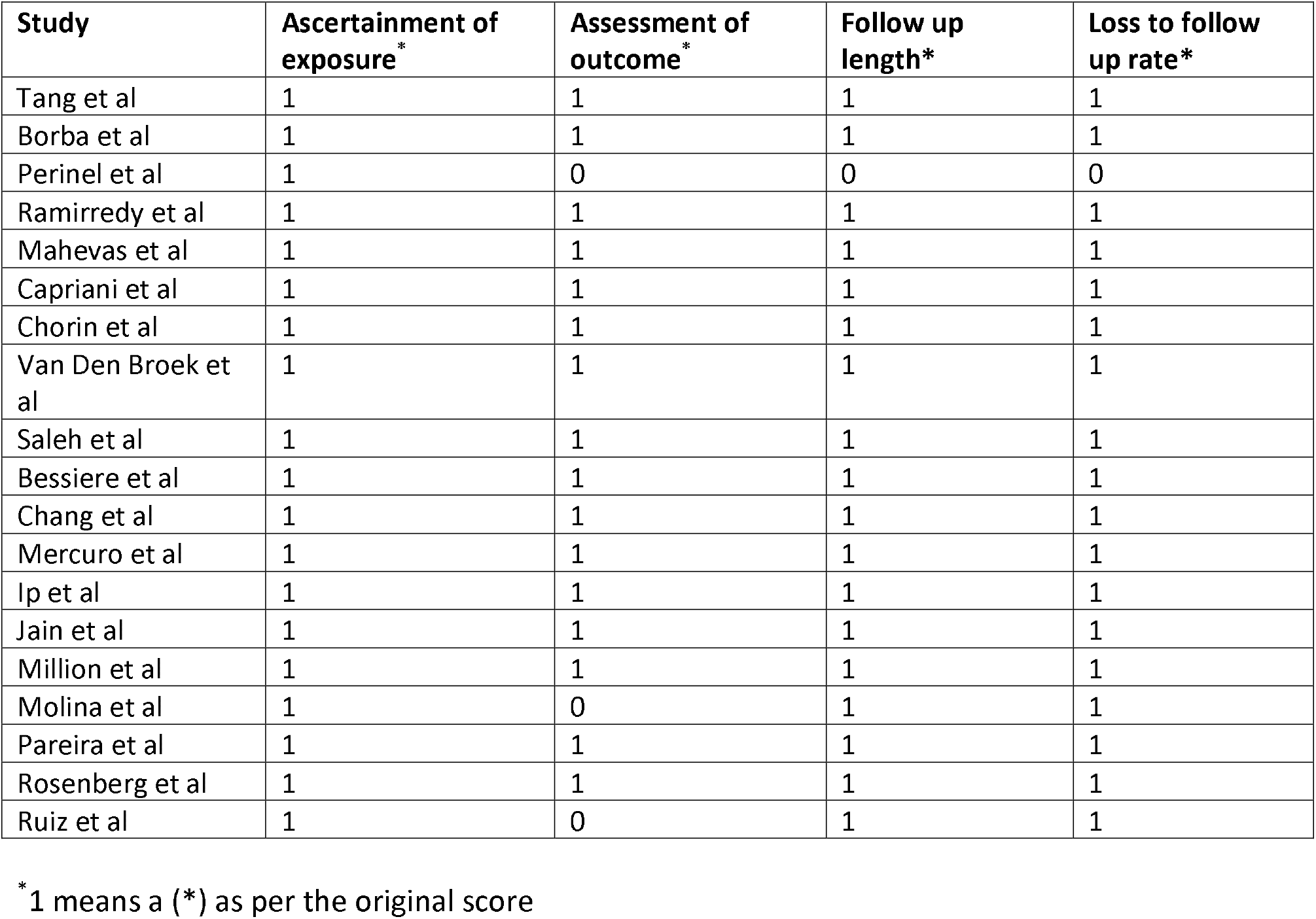
Modified Newcastle-Ottawa Quality Assessment Score

**Figure 1.**
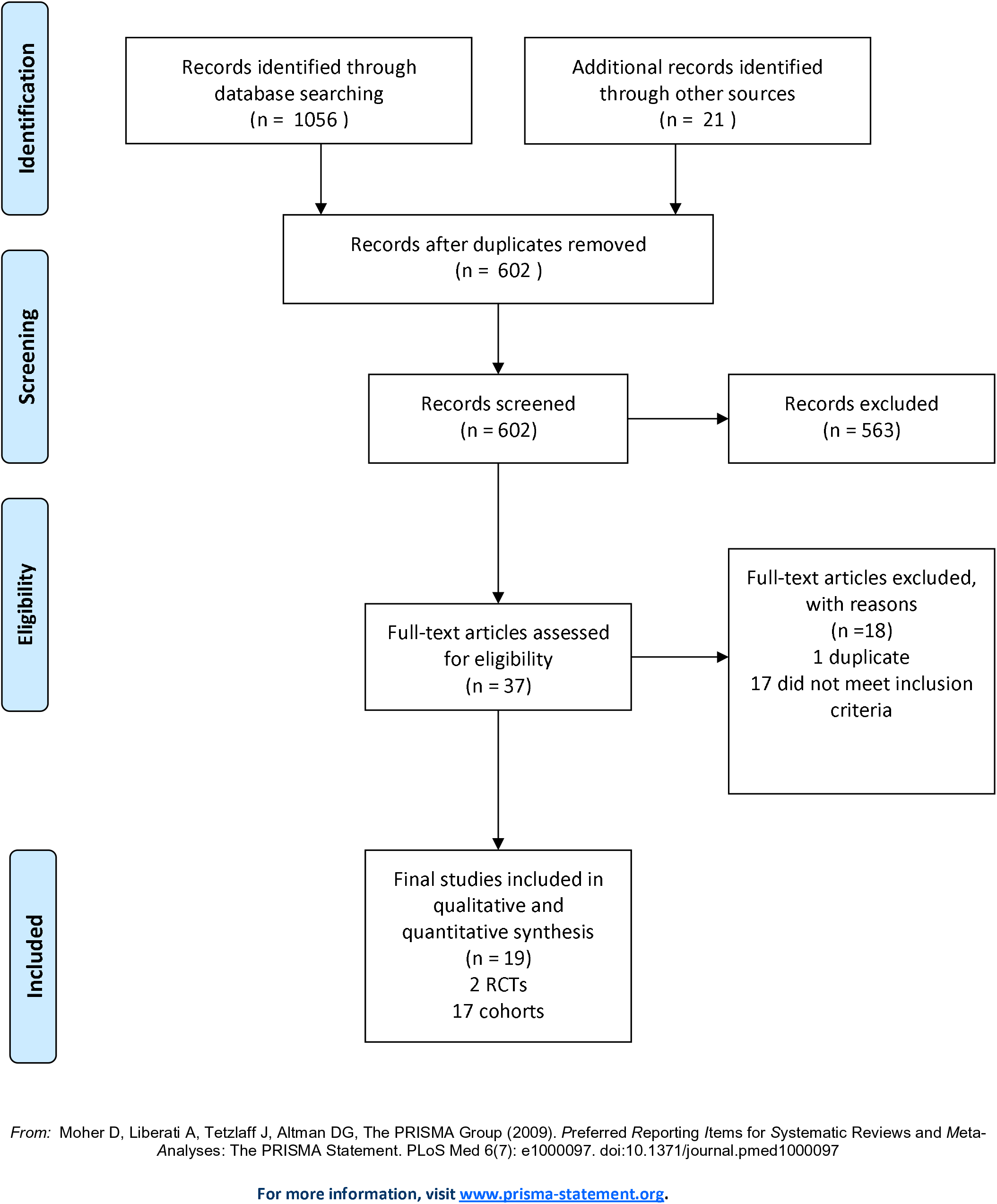
PRISMA Flow Diagram of Eligible Studies.

Rosenberg et al reported incidence of any arrhythmia (unspecified) in 19.3% and cardiac arrest in 15% in their cohort of 1006 patients who received HCQ alone or in combination with azithromycin. Non-sustained ventricular tachycardia was reported by 6 studies and was encountered in 0-5% of patients treated with CQ or HCQ. On the other hand, sustained ventricular tachycardia was described by 9 studies in 0-2.7% of patients. Only two investigators reported about new onset atrial fibrillation; it was detected in 12.8% of patients by Chang et al^49^, and in 8.5% by Saleh et al^38^.

Four studies reported on conduction abnormalities, which developed in 1-3.4% of their patients. Moreover, two studies observed acute cardiac injury defined as elevated troponin levels in 27.8%^23^, and as elevated cardiac specific creatine phosphokinase (CKMB) in 31.8%^34^. In the last study, it was noted that the CKMB elevation was higher in the high CQ dose in comparison with the lower dose (50% vs. 31.6%)^34^. Acute MI was reported by two studies. Ramiderry et al observed acute MI in 17% of their patients^47^ and Mercuro et al identified acute MI in one patient of their cohort of 90 patients^23^. Acute myocarditis was observed by Saleh et al in 1 patient (0.5%)^38^ and Borba et al in 2 patients (8.6%)^34^.

### Meta-analysis Results

TdP tachycardia, ventricular tachycardia and cardiac arrest events were observed in 156 patients in 17 studies with total of 3725 patients. The pooled incidence of these events was 3 per 1000, 95% CI (0-21), I^2^=96% (Figure 2). However, only two episodes of TdP tachycardia were reported among 2719 patients. The pooled incidence of discontinuation of CQ or HCQ due to prolonged QTc or arrhythmias was 5%, 95% CI (1-11), I^2^=98% among 4334 patients from 13 studies (Figure 3). In 12 studies of 2008 patients, the pooled incidence of change in QTc from baseline of ≥ 60 ms was 7%, 95% CI (3-14), I^2^=94% (Figure 4). Further, the pooled incidence of QTc ≥ 500 ms was 6%, 95% CI (2-12), I^2^=95% from 16 studies with 2317 patients, (Figure 5), and Among 11 studies with 3127 patients, the pooled incidence of change in QTc from baseline of ≥ 60 ms or QTc ≥ 500 ms was 9%, 95% CI (3-17), I^2^=97% (Figure 6).

**Figure 2.**
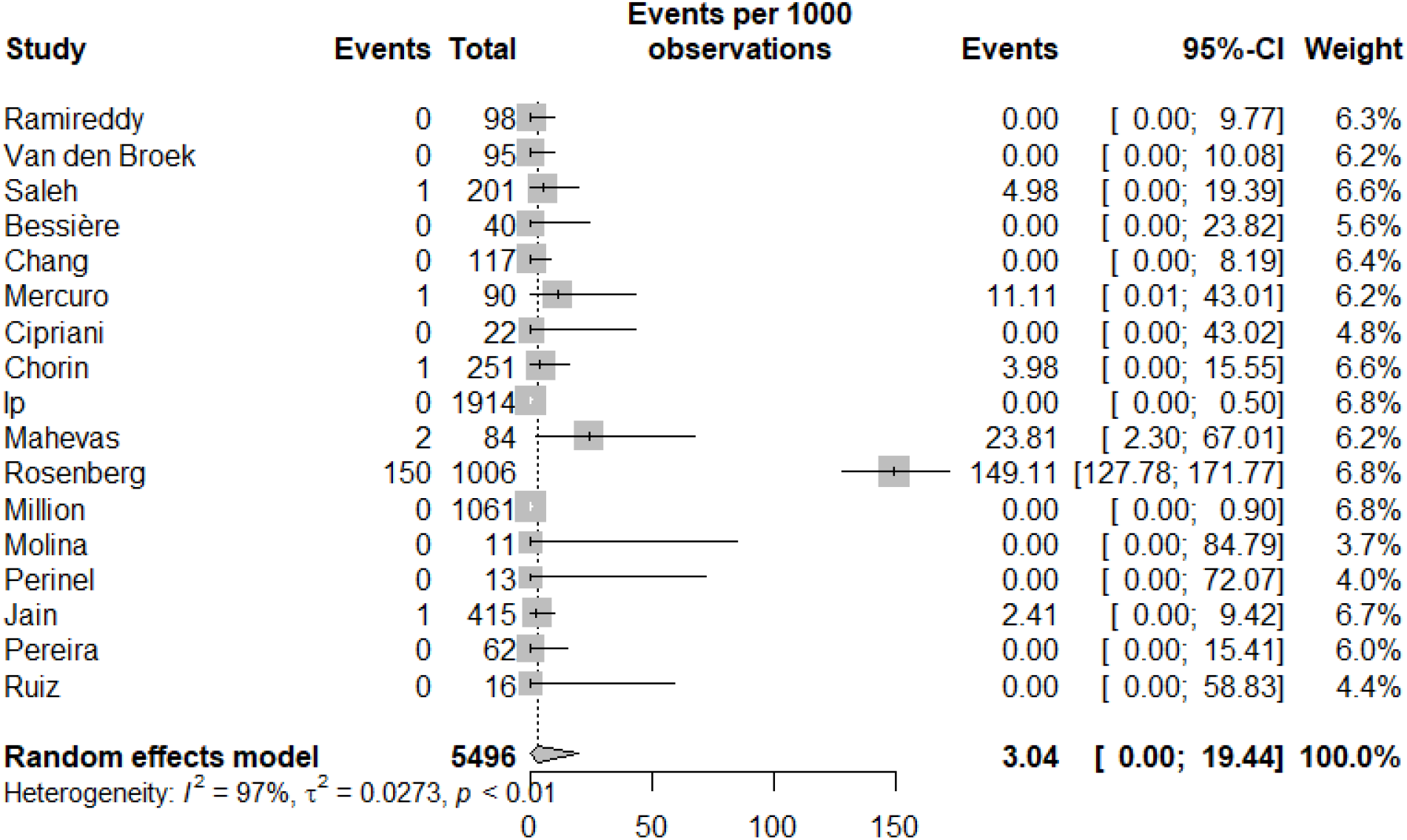
Forest plot of the pooled incidence of (torsade de pointes or ventricular tachycardia or cardiac arrest)

**Figure 3.**
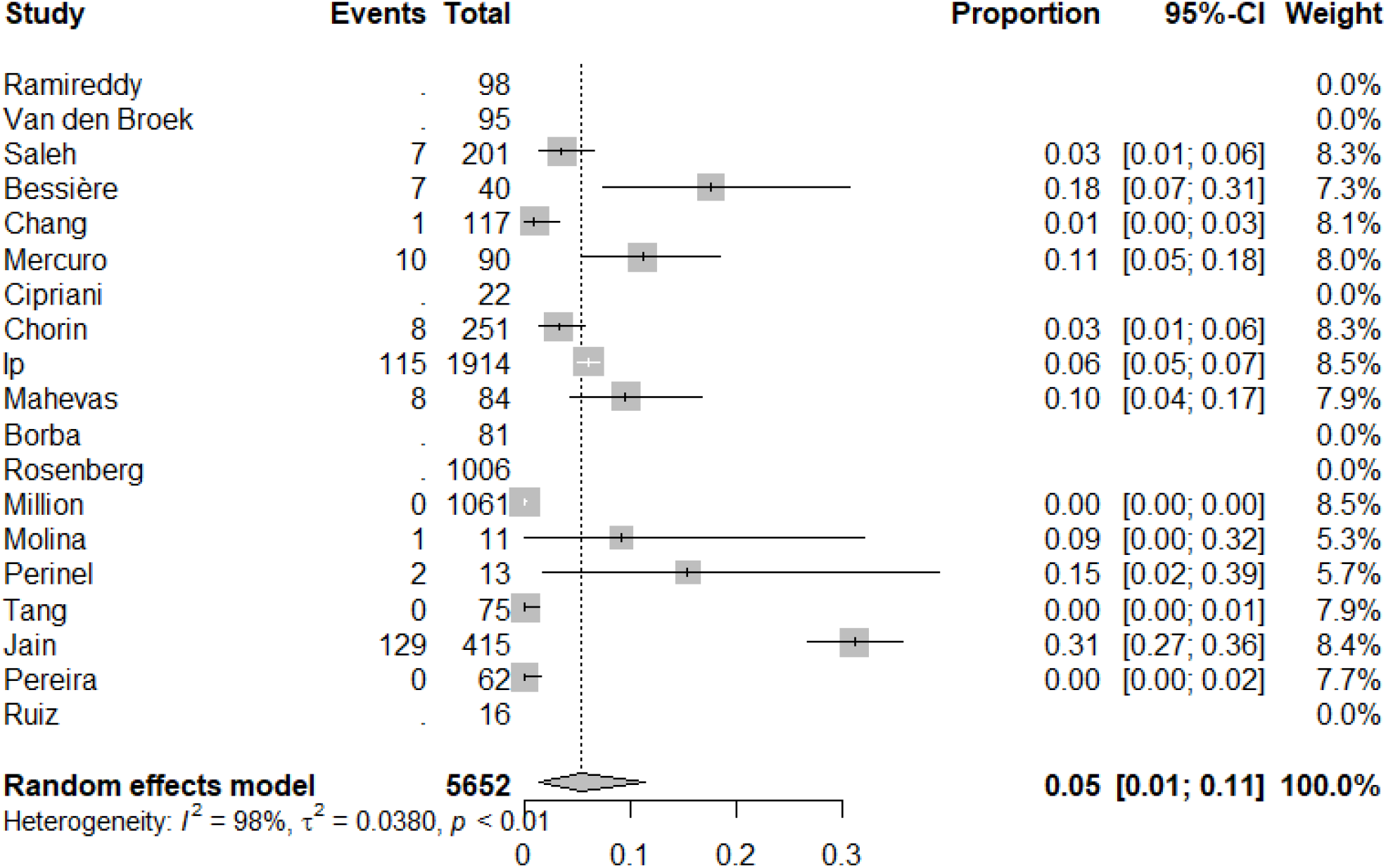
Forest plot of the pooled incidence of discontinuation of chloroquine or hydroxychloroquine due to prolonged QTc or arrythmias

**Figure 4.**
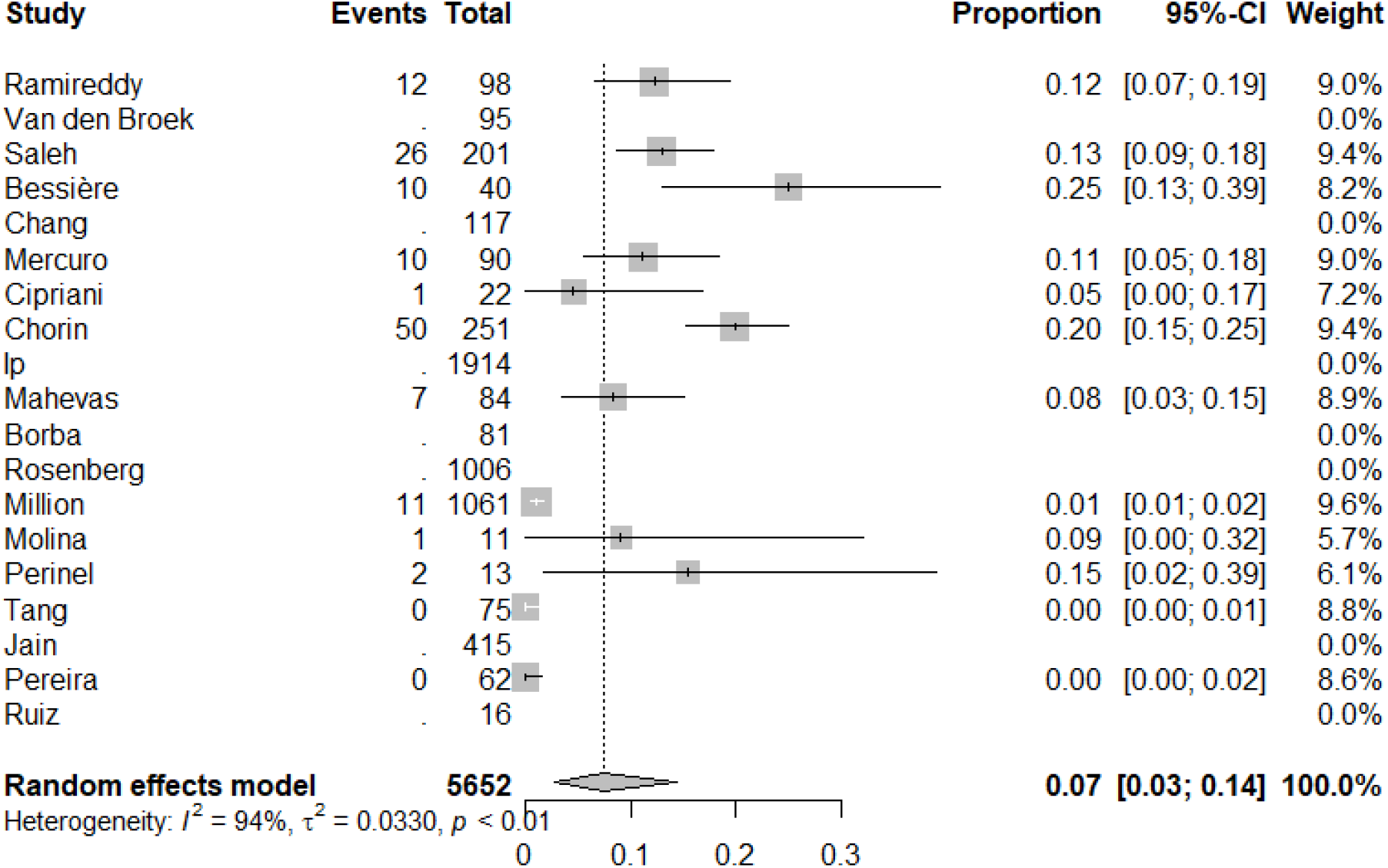
Forest plot of the pooled incidence of change in QTc from baseline of ≥ 60 ms

**Figure 5.**
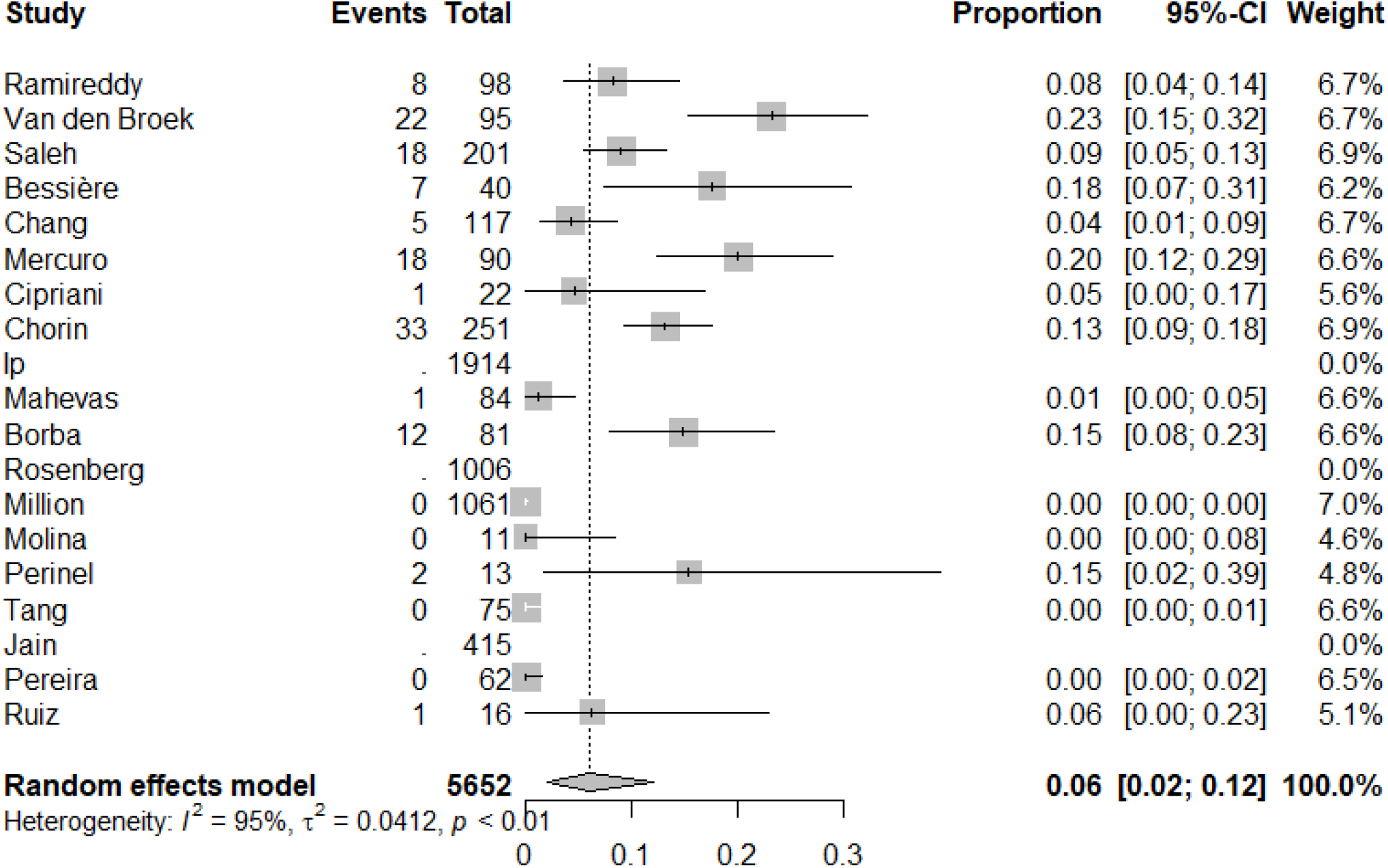
Forest plot of the pooled incidence of QTc ≥ 500 ms

**Figure 6.**
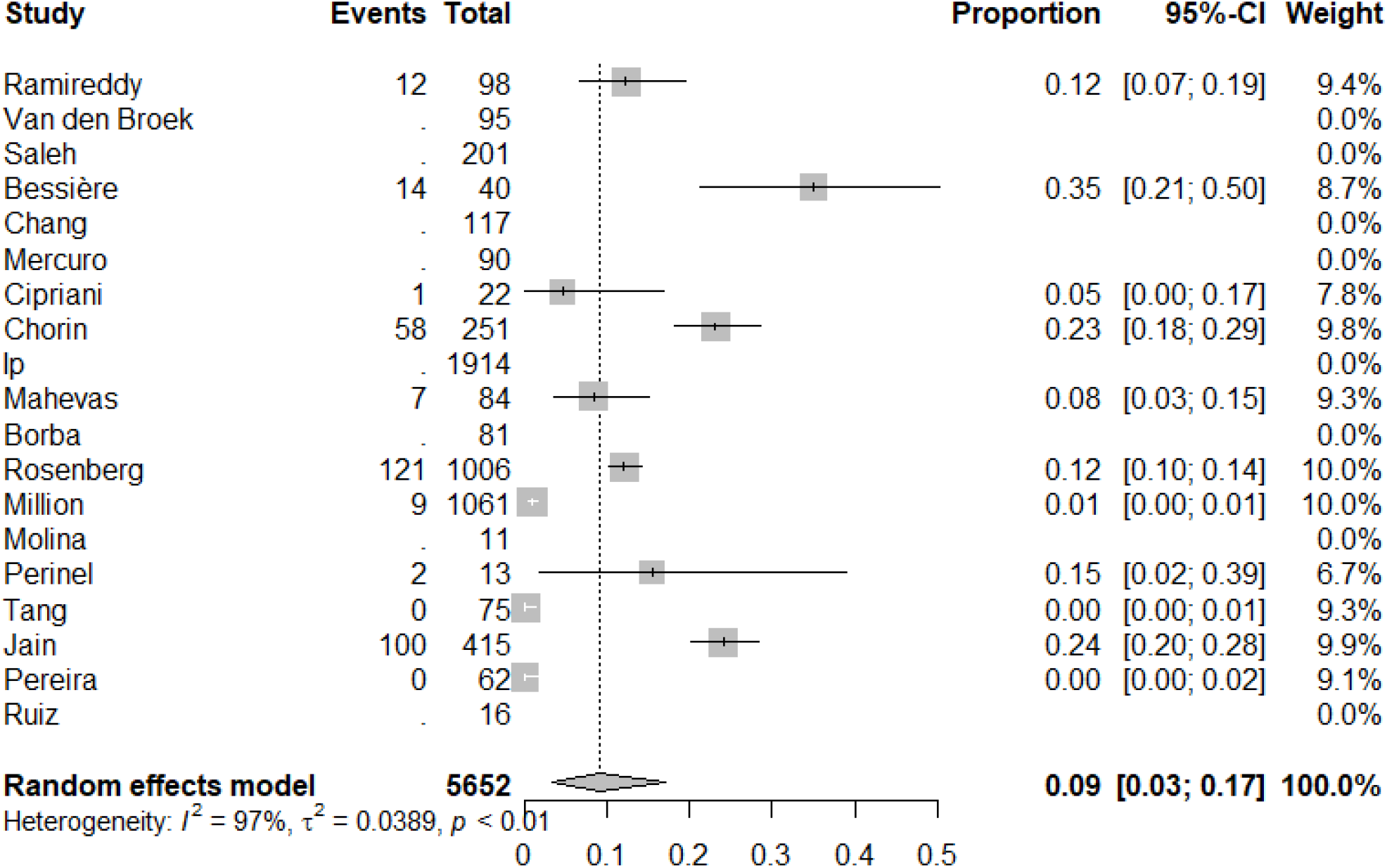
Forest plot of the pooled incidence of change in QTc from baseline of ≥ 60 ms or QTc ≥ 500 ms

### Exploring Heterogeneity

#### TdP arrhythmia or VT or cardiac arrest

Concomitant QT prolonging medications prevalence in included studies explained heterogeneity. The higher the prevalence of concomitant use of these medications, the higher the reported incidence of TdP arrhythmia or VT or cardiac arrest. The amount of heterogeneity accounted for concomitant QT prolonging medications was: 90%.

#### Discontinuation due to prolonged QTc or arrhythmias

Mean/median age, CAD, and DM prevalence in included studies explained heterogeneity. For those studies with a higher mean/median age of the study population and higher prevalence of these comorbidities, there was a higher reported incidence of discontinuation of drug due to prolonged QTc or arrhythmias. The amount of heterogeneity accounted for by age, CAD and DM was: 40%, 67% and 61%, respectively.

#### Change in QTc from baseline of ≥ 60 ms

Mean/median age, CAD, and HTN prevalence and frequency of ICU admission in included studies explained heterogeneity. The higher the mean/median age of the study population and the higher the prevalence of these comorbidities and ICU admissions, the higher the reported incidence of change in QTc from baseline of ≥ 60 ms. The amount of heterogeneity accounted for by age, CAD, HTN and ICU care was: 73%, 82%, 54% and 53%, respectively.

#### QTc ≥ 500 ms

Mean/median age, CAD, and HTN prevalence and severity of illness in included studies explained heterogeneity. The higher the mean/median age of the study population and the higher the prevalence of these comorbidities and severity of illness, the higher the reported incidence of QTc ≥ 500 ms. The amount of heterogeneity accounted for by mean/median age, and CAD, and HTN prevalence and severity of illness was: 68%, 65%, 68% and 63%, respectively.

#### Change in QTc from baseline of ≥ 60 ms or QTc ≥ 500 ms

Mean/median age, CAD, HTN, concomitant QT prolonging medications, and ICU care prevalence and severity of illness in included studies explained heterogeneity. The higher the mean/median age of the study population and the higher the prevalence of these comorbidities or use of these medications and severity of illness/ICU care, the higher the reported incidence of QTc from baseline of ≥ 60 ms or QTc ≥ 500 ms. The amount of heterogeneity accounted for mean/median age, and CAD, HTN, concomitant QT prolonging medications and ICU care prevalence and severity of illness was: 80%, 85%, 61%, 58%, 66% and 58%, respectively.

### Sensitivity Analysis

After excluding the 2 studies that used high dose CQ or HCQ^33,34^, we did not observe an important change in pooled estimates or associated heterogeneity.

## Discussion

### Findings

In this meta-analysis, we systematically examined the risk of QTc prolongation and its associated complications in patients with COVID-19 treated with antimalarial medications CQ and HCQ (CQ/HCQ). The majority of studies included in this analysis used HCQ alone or in combination with azithromycin. Our analysis revealed that treatment with CQ/HCQ was associated with a clinically significant increased risk of QTc prolongation and discontinuation of drug due to this risk. In addition, CQ/HCQ was associated with a clinically significant risk of TdP or VT or cardiac arrest of 3 per 1000 (95% CI 0.0-21).

The included studies described QTc prolongation in several ways. The reported parameters included baseline (before drug) and peak (on drug) QTc values, Δ QTc, proportion of patients with QTc ≥ 500 ms, proportion of patients with Δ QTc ≥ 60 ms, and combinations of these. The incidence of critical QTc prolongation defined as QTc ≥ 500 ms or ΔQTc ≥ 60 ms ranged from 0% to 36%. One of the most remarkable findings is that in the study by Bessière et al, 93% of the studied 40 patients exhibited an increase in QTc prolongation and 36% had critical QTc prolongation^39^. In our pooled analysis, critical QTc prolongation ranged between 6% and 9% with significant heterogeneity among the studies (I^2^ of up to 98%). Several factors contributed to the observed heterogeneity were identified by the meta-regression analysis. The factors that contributed most to the observed heterogeneity include age, hypertension, CAD, ICU admission, DM, use of other QTc prolonging agents, and COVID-19 disease severity. These factors are concordant with the biological explanation for the observed differences, as it is well known that underlying cardiac conditions, comorbidities, and inflammatory states increase the risk of drug induced QTc prolongation^50^. Severe COVID-19 disease and admission to ICU is usually associated with hypoxemia, hypotension, and electrolyte imbalance; all of which increase the risk of QTc prolongation in response to QT prolonging drugs^50-53^. These patients also manifest fever and elevated interleukin-6, which have been shown to increase the risk of QT prolongation in response to drugs and inflammation^54,55^.

This meta-analysis revealed low but clinically significant risk of combined endpoint of TdP, ventricular tachycardia and cardiac arrest. However, we could not perform a meta-analysis on TdP separately because there were only two reported cases of TdP among 2719 patients from 16 studies (0.073%). The low incidence of TdP is probably an underestimate and not applicable to other settings where ECG monitoring is not performed routinely. A number of factors can explain this low incidence of TdP; most importantly the precautionary discontinuation of the drugs when QTc reaches a certain threshold (QTc ≥ 500 ms or ΔQTc ≥ 60 ms), short duration of therapy, and in certain instances, the therapeutic intervention for long QT using QT shortening agents as reported by Saleh et al for example^38^. Indeed, in our pooled analysis, 5% of patients had their medication discontinued because of QTc prolongation and in one study by Jain et al, 30% of patients had their CQ/HCQ discontinued because of QTc prolongation^41^. Moreover, some TdP cases could have been missed because of underreporting or misclassification. In fact, the two largest studies included in this meta-analysis did not specifically include TdP as a separate endpoint but grouped all arrhythmias under one category. The study by Rosenberg et al observed arrhythmias in 19.3% and cardiac arrest in 15% of patients and it is quite possible that some of these arrhythmias were TdP or some of the cardiac arrests were preceded by TdP^44^.

Nonetheless, the incidence of TdP reported here are consistent with the published data on the drug induced TdP. The risk of developing TdP in association with non-antiarrhythmic drugs is relatively low; for instance the risk for cisapride, which has been removed from the market, was 0.001% ^56^. The risk of TdP with other non-antiarrhythmic drugs is in the range of that was reported with cisapride. The observed incidence of TdP observed in this meta-analysis (0.073%) is 73-fold higher than that of cisapride. It is noteworthy that cisapride and terfenadine (a non-sedating antihistamine) were taken off the market because the risk of TdP even though the risk of TdP associated with their use in absolute terms was very low. This underscores the importance of taking into account the total number of potential lethal events rather than the expressed ratios when assessing the risk drug induced arrhythmias. It is also well known that the highest risk for drug-induced QT prolongation and TdP is associated with class-III antiarrhythmic drugs, which ranges between 1-3% over 1-2 years^57^. The risk of TdP with Sotalol therapy at a low daily dose of 80 mg is only 0.3%^58^. This risk is much higher than the observed risk with CQ and HCQ in this study; however, the estimated risk reported for the antiarrhythmic drugs was over 1 to 2 years of chronic use as opposed to the risk reported here for CQ and HCQ over a very short-term use. In fact, this increases further the concern about the cardiac risk associated with CQ and HCQ treatment in COVID-19 disease.

Proarrhythmic drugs can also precipitate other serious arrhythmias beside TdP through reentry circuits or increased automaticity^58^ and therefore, CQ and HCQ may not be only associated with the risk of QT prolongation and TdP but they could also increase the risk of other arrhythmias. Although not all studies in this meta-analysis systematically looked for or reported about all arrhythmias, several serious arrhythmic events were reported. Four investigators reported sustained ventricular tachycardia in 5 patients. Interestingly, the 2 patients with lethal ventricular tachycardia reported by Borba et al were treated with high dose CQ^34^. In addition, Rosenberg reported QT prolongation in 12%, and arrhythmias (unspecified) and cardiac arrest in a significant number of their patients. In this study 22.5% of the total study population needed ICU admission reflecting higher disease severity with the potential increased risk of QT prolongation associated TdP and cardiac arrest^44^. The findings of QTIP study that examined the prevalence of QT prolongation among 154 acutely ill patients, confirms that QT prolongation is not only associated with TdP but also with cardiac arrest^51^. In this study, 16 patients had cardiac arrest but only one of them had TdP^51^. Other arrhythmias observed in the studies included in this meta-analysis were non-sustained ventricular tachycardia, new onset atrial fibrillation and conduction abnormalities.

A number of other cardiac adverse events documented in the included studies were not negligible and include, myocardial injury, acute MI, myocarditis, and others. Notwithstanding, a cause and effect relationship between CQ/HCQ exposure and these complications cannot be inferred from these studies. However, it is noteworthy to mention that in the study by Borba et al, the incidence of acute cardiac injury was higher in the high dose CQ group in comparison with low dose CQ (50% vs. 31.6%) and the two patients with sustained ventricular tachycardia also occurred in the high dose CQ, which could imply dose-response relationship and probable cause and effect link^34^. Additionally, the prevalence of cardiovascular comorbidities such as hypertension, CAD, heart failure, DM, atrial fibrillation, and other chronic medical conditions like chronic pulmonary and kidney disease were common across all of these studies. These observations of high prevalence of comorbidities and cardiac complications in the studies included in our meta-analysis mirror the observation in several other COVID-19 epidemiological studies ^14,15,17-21,59-61^. Prior to COVID-19, data regarding CQ/HCQ’s cardiac toxicity were scarce. One recent study looked at the Food and Drug Administration’s Adverse Event Reporting System and identified 351 TdP/QT prolongation reports linked to CQ and HCQ and almost in all of these cases these agents were used alone and not in combination with azithromycin. The relative reporting ratio (RRR) for TdP/ QT prolongation was 1.43 (CI 1.29-1.59) for CQ/HCQ and 3.77 (CI 1.80-7.87) for the combination of CQ or HCQ and azithromycin^62^. In another meta-analysis of HCQ effects on cardiovascular system, ventricular hypertrophy was noted in 22% of patients while heart failure was noted in 26.8% of patients^63^.

Another interesting observation in our meta-analysis is the common use of other QT prolonging agents in the included studies. Among the 10 studies that reported on the concomitant use of other QT prolonging agents, two of them excluded patients using other QT prolonging agents from their studies^38,49^. The concomitant use of other QT prolonging agents in the other 8 studies ranged between 24% and 100%.

It is worth noting also that it has been a common practice to use HCQ in combination with azithromycin for COVID-19 during the current pandemic. Azithromycin has been identified as a potential cause of significant serious cardiac arrhythmias through QT prolongation dependent and independent mechanisms and has been linked to increased risk of sudden cardiac death^64,65^. Hence, the concomitant use of CQ/HCQ and azithromycin or other QT prolonging agents could potentially increase the risk of serious cardiac arrhythmias and death particularly in critically ill patients or those with risk factors for QT prolongation. This contention is supported by a large observational study of 212,016 ICU patients, of whom, 6,125 were receiving QT prolonging agents. The co-administration of QT prolonging agents was associated with higher mortality and longer ICU length of stay^66^.

### Mechanisms

The principal mechanism responsible for QT prolongation produced by CQ and HCQ is due to blocking of the *KCNH2*-encoded hERG/Kv11.1 potassium channel, which is responsible for the rapidly-activating potassium current (I_Kr_). This rapidly-activating current along with the slowly-activating current (I_Ks_) are responsible for pumping potassium ions outside cardiomyocytes. The outward potassium current through these channels after the plateau phase (phase 2) of the action potential results in rapid repolarization phase of the action potential (phase 3)^67^. Genetic mutations in the genes encoding these potassium channels as well as drug-blockage of hERG/Kv11.1 potassium channels result in prolongation of the cardiac action potential that manifests as prolonged QT interval on the surface electrocardiogram ^67^. The intracellular face of the hERG channel is large and is lined with a number of aromatic residues allowing drugs like CQ and HCQ and others to bind this part of the channel and block the outward potassium current^57,67^. In a recent study using ex-vivo guinea pig and rabbit heart model, it was also shown that HCQ at the upper therapeutic concentration resulted in the generation of repolarization alternans and precipitated polymorphic ventricular tachycardia^68^.

It is also important to take in consideration that the prevalence of congenital long QT syndrome (LQTS) is approximately1 in 2000 people worldwide. Importanly, many individuals with LQTS have normal baseline QTc values, but their risk of drug-induced QT prolongation and lethal arrhythmias is increased substantially. Furthermore, 8% of individuals of African descent (p.Ser1103Tyr-SCN5A) and 2% of individuals of European descent (p.Asp85Asn-KCNE1) possess potentially pro-arrhythmic common variants associated with an increased risk of drug-induced long QT and sudden cardiac death^69^. Although the percentage of people with an inherent genetic risk is small (roughly 10%), the fact that SARS-CoV-2 infection has been spreading widely across the globe affecting millions of people, the total number of individuals at risk for drug-induced QT prolongation and TdP associated with indiscriminate use of CQ and HCQ may be unacceptably high^70^.

A recent study examined the prescription pattern of several drugs in USA and observed an almost 2000% increase in prescriptions for CQ and HCQ for fewer than 28 tablet fills for the week of March 15-21, 2020 in comparison with the same week in 2019. This surge remained steady during the following weeks^71^. This remarkable surge in CQ/HCQ use could lead to a substantial increase in preventable serious cardiac adverse events and mortality.

HCQ use has also been associated with bradycardia. In a study of mouse atria, spontaneous beating was significantly reduced by HCQ. Similarly, these findings were confirmed in sinoatrial node cells from pigs with a clear dose-dependent effect^72^. This is important for patients who might be taking HCQ and concomitant beta-blockers or amiodarone. These combinations especially in the setting of electrolyte imbalances will result in significant reduction of the automaticity of the heart and may lead to significant bradycardia. Finally, these drugs have active metabolites and relatively long elimination half-lives, especially in critically ill patients with multi-organ failure and this might increase their arrhythmogenic risk ^73^.

These observations discussed above along with our findings and the high prevalence of cardiac comorbidities and cardiovascular complication observed in COVID-19 patients indicate that the cardiac risk imposed by CQ/HCQ use in COVID-19 disease is not trivial and support the need for close monitoring of COVID-19 patients who are treated with CQ/HCQ alone or in combination with azithromycin or other QT prolonging agents. Another important aspect is the lack of strong evidence for their efficacy in improving the outcomes of COVID-19 patients and therefore, these agents should be used only in the context of randomized clinical trials given the potential harm that could be associated with their widespread use. This position is supported by the recent FDA statement^74^.

### Strength and Limitations

Our meta-analysis is the first comprehensive systematic review examining the risk of QT prolongation and its associated adverse events in COVID-19 patients treated with CQ/HCQ. However, like any meta-analysis, it has several limitations. First, the retrospective nature of most of the included studies make them prone to incomplete or missing data. Second, there were variations in the variables collected by individual studies particularly related to reporting QTc parameters and adverse events and significant differences in the patient populations enrolled by these studies. This may have resulted in underreporting of certain important endpoints like TdP. For example, the study by Rosenberg et al listed all arrhythmias under one group with a frequency of 19.3%. This study also reported high incidence of cardiac arrest of 15%, hence, it’s very possible that some cases with TdP would have been included in the arrhythmia or cardiac arrest groups. Third, there was marked heterogeneity in our pooled estimates; however, we performed a meta-regression that allowed us to identify contributors to the observed heterogeneity and further determine populations at risk for CQ/HCQ induced QT prolongation, which further strengthens our study and its conclusions.

### Implications for Ongoing Randomized Clinical Trials

A recent randomized, double-blind, placebo-controlled trial across the United States and parts of Canada tested HCQ as post-exposure prophylaxis. After high-risk or moderate-risk exposure to COVID-19, HCQ did not prevent illness compatible with COVID-19 or confirmed infection when used as post-exposure prophylaxis within 4 days after exposure^75^. Another large trial^76^, a randomized clinical trial is testing a range of potential drugs for COVID-19, including HCQ. This trial has enrolled over 11,000 patients from 175 NHS hospitals in the UK. Recently, its independent Data Monitoring Committee has reviewed the emerging data. A total of 1542 patients were randomized to HCQ and compared with 3132 patients randomized to usual care alone. There was no significant difference in the primary endpoint of 28-day mortality (25.7% HCQ vs. 23.5% usual care; hazard ratio 1.11 [95% confidence interval 0.98-1.26]; p=0.10). There was also no evidence of beneficial effects on hospital stay duration or other outcomes.

There are 104 ongoing randomized trials on HCQ in COVID-19 actively recruiting patients across different countries and continents (Supplement 2). Despite the FDA cautionary statement warning against outpatient treatment of COVID-19 patients with antimalarial, 12 trials being conducted in the US, are recruiting outpatients with COVID-19 (Supplement 3). Although all of the US trials exclude patients with known history of LQTS or arrhythmias and the majority exclude patients on other QT prolonging agents, many of them did not exclude patients with history of structural heart disease and the majority did not have adequate electrocardiographic monitoring as part of their protocol.

In the light of our findings and lack of efficacy of HCQ observed in the two RCTs discussed above, the ongoing trials should consider amending their protocols to include adequate safety monitoring and to have selective inclusion and exclusion criteria.

## Conclusion

Our meta-analysis indicates that the treatment of COVID-19 patients with CQ or HCQ alone or in combination with azithromycin is associated with a significant risk of drug-induced QT prolongation, which is a harbinger for TdP. CQ/HCQ use resulted in a relatively higher incidence of TdP as compared to drugs withdrawn from the market for this particular adverse effect. Therefore, these agents should be used only in the context of randomized clinical trials in patients at low risk for drug-induced QT prolongation and with selective inclusion and exclusion criteria and adequate safety monitoring.

## Data Availability

All data referred to in the manuscript has been provided.

## Acknowledgement

We thank Ms. Leslie Hassett, M.L.S., Librarian, at Mayo Clinic Libraries for her help with the literature search.

